# Spirituality, quality of life, and general health: a cross-sectional study

**DOI:** 10.1101/2023.05.12.23289909

**Authors:** Takeshi Yoshizawa, Abdelrahman M Makram, Sadako Nakamura, Randa Elsheikh, Engy Mohamed Makram, Nguyen Tien Huy, Kazuhiko Moji

## Abstract

**Background:** Some researchers have highlighted the need to integrate spirituality into the definition of health, as they have found a positive relationship between spirituality and quality of life (QoL). Hence, we aim to ascertain the effect of spirituality on mental health and QoL in older people residing in Kumejima Town in Japan.

**Methods:** We conducted an interview-based survey between September 2010 and 2011 on people residing in Kumejima Town aged 65 years or older. The scale used was The Spirituality Health Scale for the Elderly. The survey includes items on basic attributes (e.g., age, sex); physical, mental, and social health; spirituality; and quality of life. The analysis was conducted on a causal structure model in which spirituality is a distinct item influencing QoL (Model 1) and a causal structure model in which spirituality poses direct and indirect effects on QoL (Model 2). For each model, we reported the goodness-of-fit in terms of the goodness-of-fit index (GFI), adjusted goodness-of-fit index (AGFI), comparative fit index (CFI), and root mean square error of approximation (RMSEA). To compare the models, we used the Akaike information criterion (AIC).

**Results:** The total number of participants was 338, of whom 72.5% were female. The average age and its standard deviation were 77.2 ± 6.4 years. We presented a comparison of the results of the analysis of Model 1 and Model 2, with the results favoring Model 2. Also, Model 2 was superior to Model 1 in terms of the AIC, where the value of the AIC is 313.67 for Model 2 compared to 330.48 for Model 1.

**Conclusion:** This study was able to show the possibility of preventing a decline in QoL until death by building up spiritual health. However, further studies are required to further investigate these outcomes in the wider population (Japanese and other nationalities) and with some focus on male participants.

## Introduction

The 1946 Constitution of the World Health Organization (WHO) was the first widely known document to include physical, mental, and social well-being in the definition of health [1]. Since then, many specialists have started wondering whether this definition of health is sufficient to provide humans with better lives [2]. This has led the WHO to expand the definition into specific pieces. For example, the Ottawa Charter of 1986 included the definition of health promotion to allow people to exercise their autonomy in their everyday decision-making (e.g., what to eat, drink, or consume). The Ottawa Charter also introduced health as a resource for living [3], which led some researchers to develop health metrics such as the productivity-adjusted life-years [4]. And till this day, public health researchers are seeking to incorporate different angles of many problems in the definition of health [5].

And this brings the idea of integrating spirituality into the definition of health when the representatives of Bahrain, Libya, and Sri Lanka proposed this to the WHO Executive Board in the WHO Fifty-second World Health Assembly of 1998. The common argument was that humans are not only a body (a chemical and physical form) but are also minds and souls. Moreover, because the definition of spirituality differs from one country to another, there was difficulty in incorporating spirituality into the definition since it was first proposed in 1984. They also emphasized that religion is not to be confused with spirituality [6]. In their systematic review conducted to define spirituality, de Brito Sena et al. summarized that spirituality includes 24 dimensions, most notably a connection/relation to something or someone, a meaning/purpose of life, believing in something divine or with limitless power like a God, leading an immaterial life, or having good community relationships [7]. Other domains are also crucial to understanding how the mind of a human shapes their experiences, even in the healthcare setting [7, 8].

To cover the aspect of linking spirituality to health, one should acknowledge that suffering without meaning is the ultimate doom of our race [9]. For example, a patient suffering from a chronic illness may question their reality and fate. They may even become reluctant to receive their prescribed medicines [10]. Similarly, some studies have found that people who practice spiritual activities may live longer [11], are less stressed [12], cope better with pain [13–15], and have a better quality of life (QoL) [16]. Other benefits also exist [8].

However, the notion that QoL declines as death approaches focuses entirely on the biological aspects of human beings. For example, previous studies with older people have demonstrated that QoL also relates to frailty, mental health, social factors, psychological elements, and the surrounding environment. This shows that physical health and QOL do not necessarily fit into a linear relationship in older people [17–21]. Furthermore, research on aging has shown that the degree of spirituality is more potent in older people who are physically frail, suggesting that even if physical health declines with old age, spirituality can provide life satisfaction and better QoL [22, 23]. Unfortunately, however, research on spirituality in Japan is dominated by qualitative studies, and there has been little validation in quantitative studies using scales to measure spirituality.

Therefore, this study was based on the hypothesis that spirituality should be included as the fourth health dimension in the WHO definition of health, i.e., physical, mental, and social health, as it influences the QoL of older people. We aimed to determine the relationship between spiritual health, mental health, and quality of life of old people.

## Methods

### Study design

The reporting of this survey study was checked against Consensus-Based Checklist for Reporting of Survey Studies (CROSS) [24], and the checklist is provided as Supplementary Table 1.

### Study setting

This study was conducted on people aged 65 years or older, who did not require nursing care, and who lived in Kumejima-cho, Okinawa Prefecture. Kumejima-cho, the surveyed area, is located on the East China Sea, 100 km west of Naha City on the main island of Okinawa, and is an island region with a circumference of approximately 48 km and an area of 63.65 km² (Figure 1). There are 3-5 daily air flights from Okinawa Naha Airport, which take approximately 25 minutes. There are one or two boat services a day, which takes about three hours.

**Figure 1.**
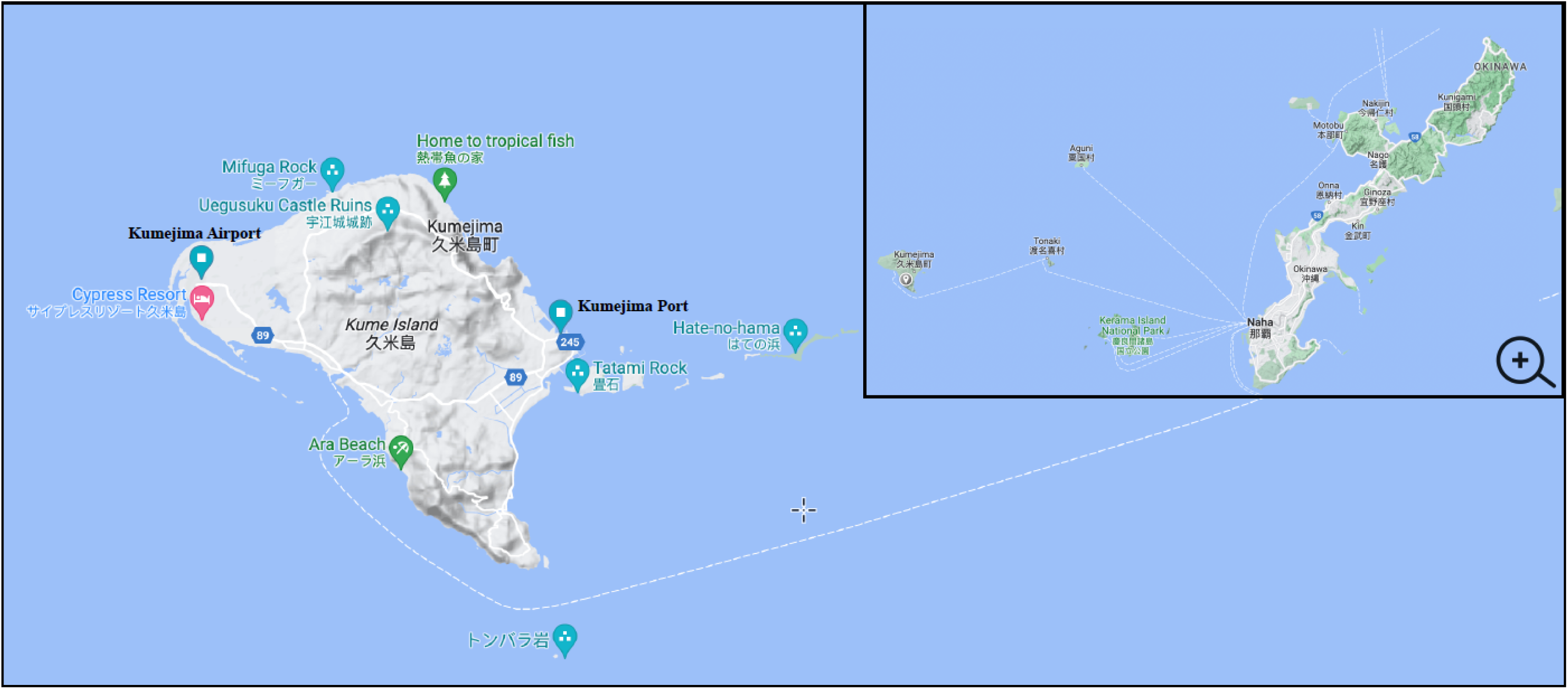
A map to illustrate the geography of the island with respect to Japan.

In 2020, Kumejima Town had a population of 7,192, slightly more male residents, a population density of 113.0/km², and a −1.5% annual change in population [25]. In Kumejima Town, the number of households with elderly persons aged 65 or older is 809, of which 311 are elderly couple-only households and 310 are elderly single-person households. The population aged 65 and over in Kumejima Town is 1,685, with an aging rate of 20.1% (2010). This is slightly higher than the rate for Okinawa Prefecture as a whole (17.5% in 2009) and the national average (22.7% in 2009) [26].

### Survey development and conduction

The survey was conducted in September 2010 and 2011 in an interview format and using a collective method. The Spirituality Health Scale for the Elderly (hereafter referred to as the SP Health Scale) [27] was selected as the scale to measure spirituality because it was developed as a scale to measure the spirituality of older people and has been tested for reliability and validity, considering the socio-cultural context of Japan. The survey was in Japanese and was already tested and validated before [27]. The survey includes items on basic attributes (e.g., age, sex); physical, mental, and social health; spirituality; and quality of life. The collected data did not include any information that could identify individual participants.

We had three questions about physical health. Although we used the same survey, we made one change: the question “Can you do personal things by yourself?” was answered using a four-question system, with “All”, “Mostly”, “Sometimes” and “Not much” as the possible answers. The mental health items were also three and were about “subjective sense of health”, “subjective sense of satisfaction”, and “sense of stress”. The social health also included three items which were about the “frequency of going out”, “degree of neighborhood interaction”, and “role in family and community”.

Spirituality was based on the SP Health Scale, which consists of six sub-concepts: “meaning and purpose of living”, “attitudes toward death and dying”, “self-transcendence”, “accordance with others”, “spiritual support”, and “harmony with nature”, with three questions for each sub-concept, making a total of 18 items.

### Data analysis

The baseline characteristics were summarized in mean, standard deviation, and range for continuous variables and event and frequency for categorical ones. The analysis was conducted on a causal structure model in which spirituality is a distinct item influencing QoL (Model 1, Figure 2A) and a causal structure model in which spirituality poses direct and indirect effects on QoL (Model 2, Figure 2B).

**Figure 2.**
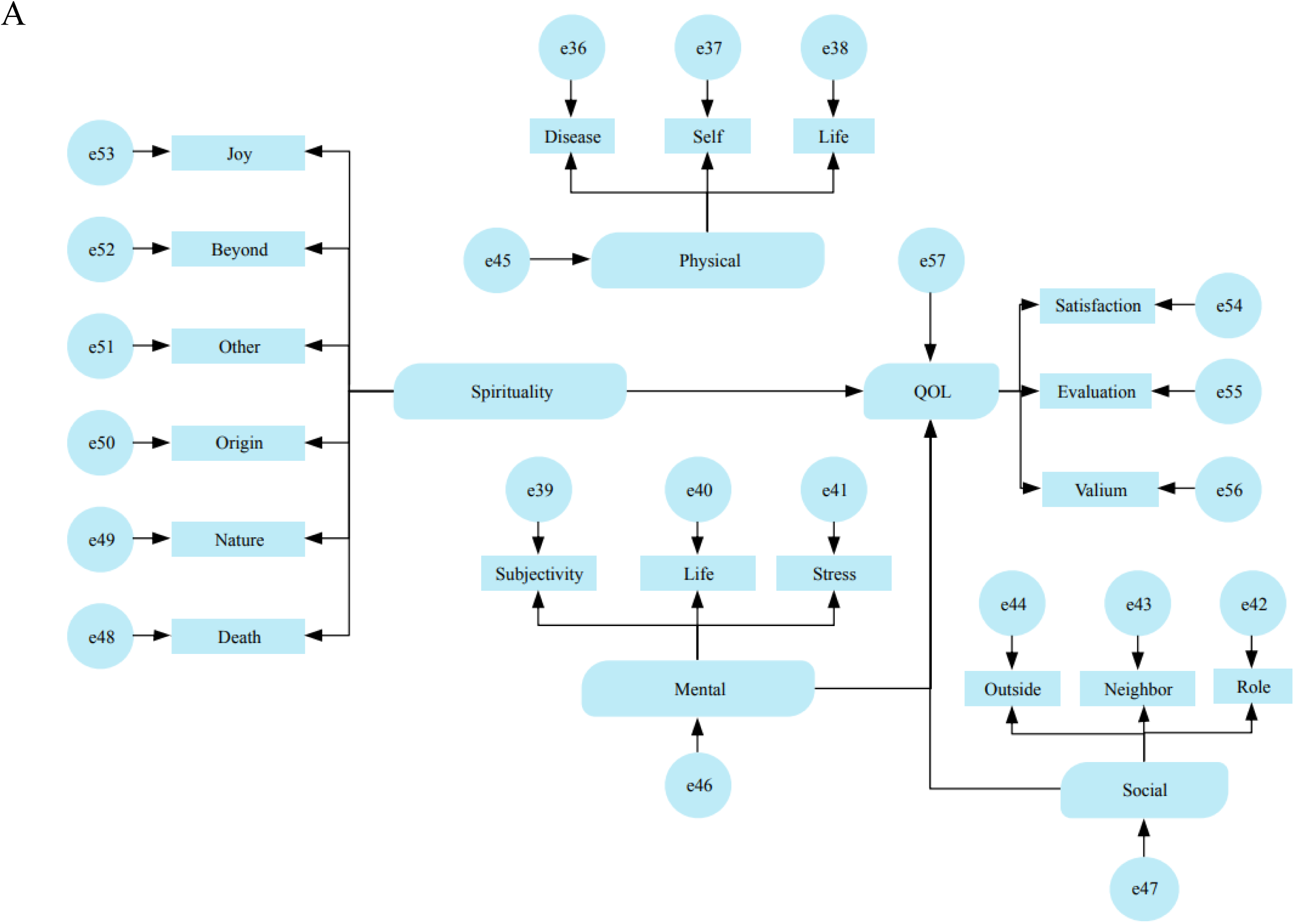
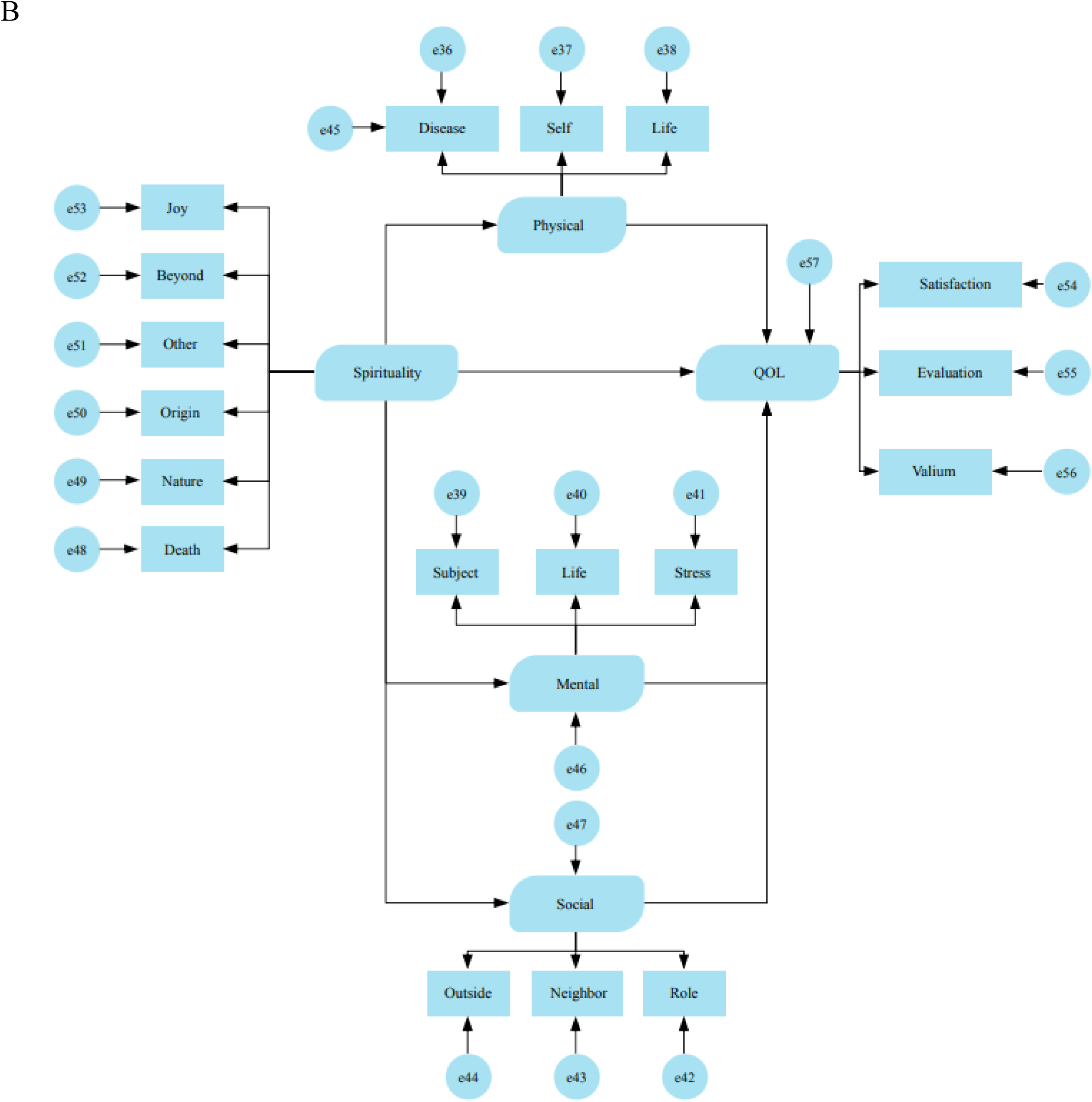
The causal structure model in which spirituality is a distinct item influencing QoL (Model 1, Figure 2A) and a causal structure model in which spirituality poses direct and indirect effects on QoL (Model 2, Figure 2B)

The covariance structure analysis was used to test the comparisons in terms of model fit, standardized path coefficients, overall effect, and coefficient of determination. For each model, we reported the goodness-of-fit in terms of the goodness-of-fit index (GFI), adjusted goodness-of-fit index (AGFI), comparative fit index (CFI), and root mean square error of approximation (RMSEA). Finally, to compare the models, we used the Akaike information criterion (AIC).

## Results

The subjects who agreed to be interviewed were 338 elderly people (93 males and 245 females). Approximately three-quarters of the respondents were women (245/338, 72.5%). The average age and its standard deviation were 77.2 ± 6.4 years. The most common family structure was living with a spouse (34.3%), followed by living with a spouse and children (30.2%), and living alone (15.4%). The most common final educational level was up to compulsory education (76.0%), up to high school (20.1%), and university or higher (3.8%). The mean height and standard deviation were 157.4 ± 6.1 cm for men and 144.8 ± 5.3 cm for women, and the mean weight and standard deviation were 59.9 ± 8.9 kg for men and 51.8 ± 8.3 kg for women. The body mass index (BMI) was 24.1 ± 3.0 kg/m^2^ for men and 24.7 ± 3.5 kg/m^2^ for women. Blood pressure was 144.0 ± 20.4 mmHg systolic in men and 145.2 ± 20.2 mmHg in women, and 77.3 ± 11.4 mmHg diastolic in men and 77.9 ± 11.2 mmHg in women (Table 1).

**Table 1.** Baseline characteristics of the sample (n=338). Continuous variables are presented in mean ± standard deviation (range)

| Category |  | Frequency | (%) |
| --- | --- | --- | --- |
| Sex | Male | 93 | 27.5% |
|  | Female | 245 | 72.5% |
| Age (years) | | 77.2±6.4 (65-94) | |
| Family structure | Single | 52 | 15.4% |
|  | Married | 116 | 34.3% |
|  | Married with children | 102 | 30.2% |
|  | Separate with children | 56 | 16.6% |
|  | Other | 12 | 3.6% |
| Education | Below high school | 257 | 76.0% |
|  | High school | 68 | 20.1% |
|  | University or above | 13 | 3.8% |
| Height (cm) | Actual value | 148.3±7.9 (129.5-174.0) | |
| | Self-reported value | 149.4±8.1 (120.0-175.0) | |
| Weight (Kg) | Actual value | 54.0±9.2 (31.5-83.6) | |
| | Self-reported Value | 53.9±8.7 (32.0-78.0) | |
| BMI (kg/m<sup>2</sup>) | Actual values | 24.5±3.4 (15.3-35.7) | |
| | Self-reported value | 23.9±3.1 (16.0-34.2) | |
| Blood pressure (mmHg) | Systolic blood pressure | 144.9±20.3 (87.0-216.0) | |
| | Diastolic blood pressure | 77.8±11.2 (42.0-115.0) | |
| Taking medication for blood pressure | Yes | 206 | 60.9% |
|  | No | 132 | 39.1% |

For Model 1, the results of the various goodness-of-fit indices are presented in Table 2. The chi-squared value was 250.480, with a probability of 0.000. No significant reduction in the chi-squared value was expected by the modification indicators: the GFI was 0.924, the AGFI was 0.900, the CFI was 0.844, and the RMSEA was 0.052. The results of the standardized path coefficients are presented in Table 3. The path coefficient for mental health to QoL was 0.588, which was significant (p<0.01). The path coefficient from social health to QoL was 0.253, which was significant (p<0.05). The overall effect on QoL (direct effect + indirect effect; however, only the direct effect was used in this model) is shown in Table 4. The largest effect was 0.588 for spiritual health to QoL, followed by 0.401 for physical health to QoL, then 0.253 for social health to QoL, and the smallest effect was 0.116 for spirituality to QoL. The coefficient of determination for QoL, the objective variable of the model, was 0.583.

**Table 2.** Main goodness-of-fit indicators for hypothesis Model 1.

| Fit indicators |  | P-value |
| --- | --- | --- |
| Chi-squared test | Chi-squared value | 250.480 |
|  | Degree of freedom | 131 |
|  | Probability | 0.000 |
| GFI |  | 0.924 |
| AGFI |  | 0.900 |
| CFI |  | 0.844 |
| RMSEA |  | 0.052 |

Abbreviations: GFI (Goodness-of-fit index); AGFI (Adjusted goodness-of-fit index); CFI (Comparative fit index); RMSEA (Root mean square error of approximation).

**Table 3.** Results of covariance structure analysis for hypothesis Model 1 (standardized path coefficients)

| Path |  |  | Path Factor | Significant difference |
| --- | --- | --- | --- | --- |
| Physical health | → | QoL | 0.401 | - |
| Mental health |  |  | 0.588 | ** |
| Social health |  |  | 0.253 | * |
| Spirituality |  |  | 0.116 | - |
| Physical health | → | Disease | 0.176 | - |
|  |  | Self-awareness | 0.676 | - |
|  |  | Life activities | 0.113 | ※1 |
| Mental health | → | Subjectivity | 0.638 | ※1 |
|  |  | Life satisfaction | 0.569 | ** |
|  |  | Stress | 0.441 | ** |
| Social health | → | Role in society | 0.423 | ※1 |
|  |  | Neighborhood | 0.502 | ** |
|  |  | Going outside | 0.467 | ** |
| Spirituality | → | Dying | 0.233 | ** |
|  |  | Nature | 0.623 | ** |
|  |  | Origin | 0.715 | ** |
|  |  | Other | 0.638 | ** |
|  |  | Go above and beyond yourself | 0.554 | ** |
|  |  | Joy of living | 0.691 | ** |
| QoL | → | Satisfaction | 0.469 | ※1 |
|  |  | Evaluation | 0.387 | ** |
|  |  | Valium | 0.533 | ** |

※1 The path to the observed variable is fixed to 1
\*\* P<0.01
\* P<0.05
Abbreviations: QoL (Quality of life).

**Table 4.** Effects on dependent variable in hypothesis Model 1 (direct and total effects)

| **Dependent variable** | **Independent variables** | **Direct effects** | **Indirect effects** | **Overall Effects** |
| --- | --- | --- | --- | --- |
| QoL | Physical health | 0.401 | - | 0.401 |
|  | Mental health | 0.588 | - | 0.588 |
|  | Social Health | 0.253 | - | 0.253 |
|  | Spirituality | 0.116 | - | 0.116 |

Abbreviations: QoL (Quality of life).

As for Model 2, the results of the various goodness-of-fit indices are presented in Table 5. The chi-squared value was 227.670, with a probability of 0.000. None of the modification indicators could be expected to significantly reduce the chi-squared value: the GFI was 0.929, the AGFI was 0.906, the CFI was 0.870, and the RMSEA was 0.048, all of which resulted in higher acceptance levels than in Model 1. The results of the standardized path coefficients are presented in Table 6. The path coefficient from spirituality to spiritual health was 0.363, which was significant (p<0.01). The path coefficient from spiritual health to quality of life was 0.622, which was significant (p<0.01). The overall effect (direct + indirect) on QoL is shown in Table 7. The largest effect was 0.622 for spiritual health to QoL, followed by 0.377 for physical health to QoL, 0.278 for spirituality to QoL, and the smallest effect was 0.240 for social health to QoL. When the overall effect of spirituality on QoL was split into direct and indirect effects, they were −0.008 and 0.286 respectively. This means that the indirect effect of spirituality on QoL is much larger than the direct effect. The coefficient of determination for QoL as the objective variable in this model was 0.610, indicating that a higher proportion was explained than in Model 1.

**Table 5.** Main goodness-of-fit indicators for hypothesis Model 2.

| Fit indicators |  | P-value |
| --- | --- | --- |
| Chi-squared test | Chi-squared value | 227.670 |
|  | Degree of freedom | 128 |
|  | Probability | 0.000 |
| GFI |  | 0.929 |
| AGFI |  | 0.906 |
| CFI |  | 0.870 |
| RMSEA |  | 0.048* |

Abbreviations: GFI (Goodness-of-fit index); AGFI (Adjusted goodness-of-fit index); CFI (Comparative fit index); RMSEA (Root mean square error of approximation).

**Table 6.**
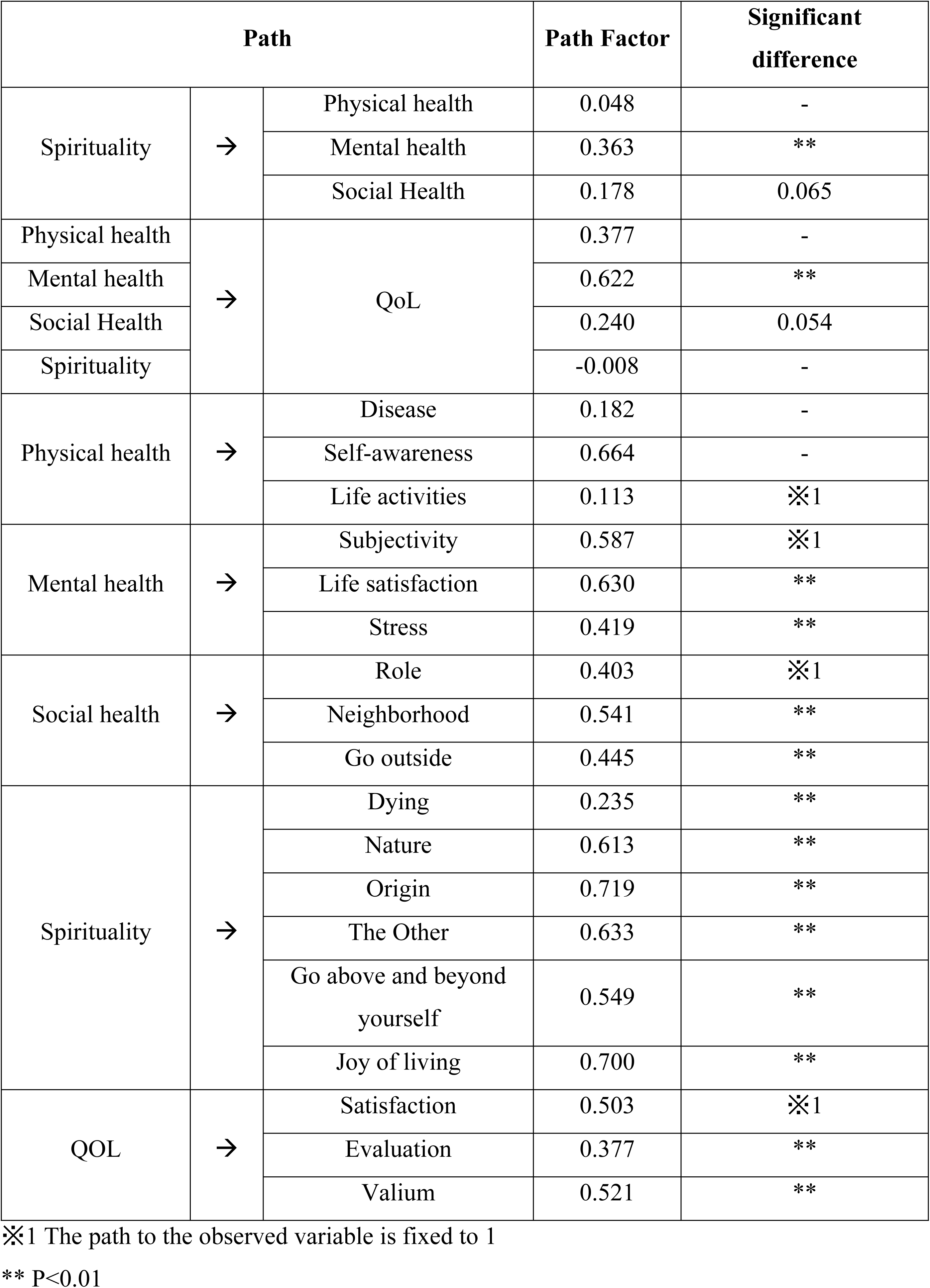
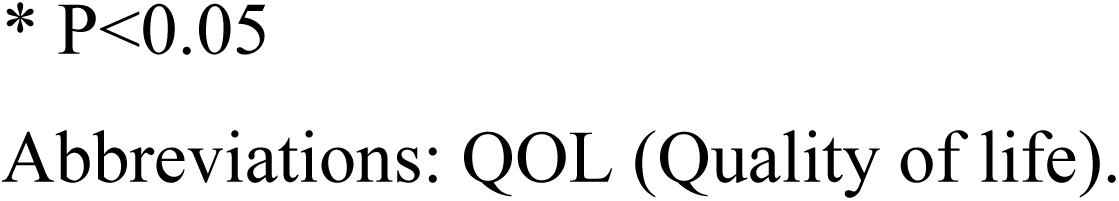
Results of covariance structure analysis for hypothesis Model 2 (standardized path coefficients)

| Path |  |  | Path Factor | Significant difference |
| --- | --- | --- | --- | --- |
| Spirituality | → | Physical health | 0.048 | - |
|  |  | Mental health | 0.363 | ** |
|  |  | Social Health | 0.178 | 0.065 |
| Physical health | → | QoL | 0.377 | - |
| Mental health |  |  | 0.622 | ** |
| Social Health |  |  | 0.240 | 0.054 |
| Spirituality |  |  | -0.008 | - |
| Physical health | → | Disease | 0.182 | - |
|  |  | Self-awareness | 0.664 | - |
|  |  | Life activities | 0.113 | ※1 |
| Mental health | → | Subjectivity | 0.587 | ※1 |
|  |  | Life satisfaction | 0.630 | ** |
|  |  | Stress | 0.419 | ** |
| Social health | → | Role | 0.403 | ※1 |
|  |  | Neighborhood | 0.541 | ** |
|  |  | Go outside | 0.445 | ** |
| Spirituality | → | Dying | 0.235 | ** |
|  |  | Nature | 0.613 | ** |
|  |  | Origin | 0.719 | ** |
|  |  | The Other | 0.633 | ** |
|  |  | Go above and beyond yourself | 0.549 | ** |
|  |  | Joy of living | 0.700 | ** |
| QOL | → | Satisfaction | 0.503 | ※1 |
|  |  | Evaluation | 0.377 | ** |
|  |  | Valium | 0.521 | ** |

※1 The path to the observed variable is fixed to 1
\*\* P<0.01
\* P<0.05
Abbreviations: QOL (Quality of life).

**Table 7.** Effects on dependent variable in hypothesis Model 2 (direct, indirect, and total effects)

| **Dependent variable** | **Independent variables** | **Direct effects** | **Indirect effects** | **Overall Effects** |
| --- | --- | --- | --- | --- |
| Physical health | Spirituality | 0.048 | - | 0.178 |
| Mental health | Spirituality | 0.363 | - | 0.363 |
| Social Health | Spirituality | 0.178 | - | 0.048 |
| QoL | Physical health | 0.377 | - | 0.377 |
|  | Mental health | 0.622 | - | 0.622 |
|  | Social Health | 0.240 | - | 0.240 |
|  | Spirituality | -0.008 | 0.286 | 0.278 |

Abbreviations: QoL (Quality of life).

A comparison of the results of the analysis of Model 1 and Model 2 is presented in Table 8. The AIC was added as an indicator to compare the models. The value of the AIC is 313.67 for Model 2 compared to 330.48 for Model 1. The relative superiority of Model 2 in terms of the coefficient of determination, goodness of fit index, and AIC is shown.

**Table 8.** Comparison of covariance structure analysis results in hypothesis Models 1 and 2 (main goodness-of-fit indicators and coefficients of determination of dependent variable)

| Model | Chi-squared test |  |  | GFI | AGFI | CFI | RMSEA | AIC | Coefficient of determination (QoL) |
| --- | --- | --- | --- | --- | --- | --- | --- | --- | --- |
|  | Chi-squared value | Degree of freedom | Probability |  |  |  |  |  |  |
| Hypothesis Model 1 | 250.480 | 131 | 0.000 | 0.924 | 0.900 | 0.844 | 0.052 | 330.480 | 0.583 |
| Hypothesis Model 2 | 227.670 | 128 | 0.000 | 0.929 | 0.906 | 0.870 | 0.048 | 313.670 | 0.610 |

Abbreviations: GFI (Goodness-of-fit index); AGFI (Adjusted goodness-of-fit index); CFI (Comparative fit index); RMSEA (Root mean square error of approximation); AIC (Akaike information criterion).

## Discussion

In this study, we aimed at determining the relationship between spiritual health, mental health, and quality of life in the older population to advocate for amending the definition of health to include spirituality. But before delving into concrete conclusions, one must consider the wider aspects of the sampled population and how they may differ in a broader context.

Our sample had a BMI of 24.1±3.0 kg/m^2^ for men and 24.7±3.5 kg/m^2^ for women, slightly higher than the mean BMI of 23.1±3.2 kg/m^2^ for men and 23.1±3.6 kg/m^2^ for women aged 70 or older in the 2010 (the time of survey conduction) National Health and Nutrition Survey [28]. Moreover, systolic blood pressure was 144.0±20.4 mmHg in men and 145.2±20.2 mmHg in women, and diastolic blood pressure was 77.3±11.4 mmHg in men and 77.9±11.2 mmHg in women, generally within the normal to borderline hypertension range for the elderly as defined by the WHO in their 1993 guidelines [29], but this is regarded as hypertension in more recent global guidelines [30]. Logically, more than half of the participants (206, 60.9%) were taking blood pressure medication. Although this result was slightly higher than the 52.7% result for the use of blood pressure-lowering medication in the National Health and Nutrition Survey [28], the analyzed population in this study was considered to be an average elderly population.

Using quantitative data, the study examined the contribution to QoL of older people regarding a health concept that includes spirituality as a fourth health dimension to physical, mental, and social health. This was proposed by previous research that a more comprehensive health model is possible by integrating aspects of physical, mental, and social health from a spiritual perspective and that the three categories of human existence are physical/material (corporeal), mental, and spiritual (spiritual) [31]. The classification of human existence into three categories: corporeal, mental, and spiritual, and the recommendation of previous research that these three are not to be equated and parallel, but that spirituality occupies a position of foundation on which corporeal and mental can be built [32].

The contribution of spirituality was found to be not a direct effect on the QoL of older people, but an indirect effect via social and mental health, with a particularly strong contribution to mental health. This is in line with Suzuki’s idea that the awakening of spirituality, which is a function that exists deep within the psyche, brings the psyche into contact with its own subject and true identity, fully manifests its original activities, and attains true significance [33].

As physical health was hardly involved in this (standardized path coefficient from spirituality to physical health: 0.048), it was considered appropriate to view it as an antagonistic relationship with spirituality. In other words, it was thought that this may be an indication that spirituality is stronger in the physically frail, which has been previously reported [34, 35]. This can be easily imagined as older people are likely to be in a crisis where they are forced to reconsider the meaning of their existence (e.g., the meaning of life) due to loss of physical functions, bereavement of family and friends, retirement from social roles, and so on. So, they tend to view their own death as an inevitable end, more realistically and routinely, and, like terminally ill cancer patients, the elderly living with old age can be seen as a population with a growing interest in spirituality.

We believe that future cohort studies will demonstrate that even if physical health declines, it is possible to maintain or improve QoL by increasing spirituality and influencing mental health. Regarding the fact that there was little direct effect between spirituality and QoL (standardized path coefficient from spirituality to QoL: −0.008), the results of a meta-analysis of studies examining the relationship between spirituality and QoL stated that although the relationship between the two was not completely unrelated, it was not necessarily strong. This may be due to the existence of some mediating factor between the two relationships [36]. This mediating factor was thought to be precisely the mental health in this study.

This study has some limitations, and some of them were previously discussed in various parts. These include the slightly different characteristics of the sample when compared to the average Japanese population. Moreover, it was not feasible to include more people due to the nature of data collection (interviews). Further, females were more likely to respond to our call for interviews and were more engaging, rendering the results of this study more representative of the female population. It is also important to highlight the fact that spirituality may be influenced by ancestral folk beliefs and that these beliefs in Kumejima town are almost identical. Therefore, future studies need to consider the effect of different religions on spirituality and QoL. Lastly, conducting this survey in urban areas may yield very different results.

## Conclusion

The study was able to show that there is an association between spiritual health, mental health, and QoL through spirituality to improve the QoL of older people. This contrasts with the conventional view that health declines with age and worsening illness, and therefore QoL gradually declines as people approach death, whereas healthy older people can be expected to continue to improve their QoL as they age, even if they become somewhat ill, experience increased subjective symptoms, have their physical health compromised, and experience an inevitable decline in their QoL. This suggests the possibility of preventing a decline in QoL and remaining happy until death by enhancing spirituality. And this opens a new door to improving QoL in adjunction with the developing medical technology. In the future, not only the elderly, but all generations may need to think of health as coexisting with illness. We believe that the concept of health with the addition of spirituality may be in line with such ideas.

## Declarations

### Ethical approval

This study was conducted under approval number 107 of the Medical Ethics Review Committee of Kagawa Nutrition University.

### Consent to participate

The objectives and content of the interview survey were explained in writing to the Kumejima Town civil workers, and their consent was obtained. The Kumejima civil workers explained the purpose of the study to the ward leaders of each village and the leaders of the Fureai Salons and obtained their consent. The Kumejima Town civil workers also explained the purpose of the research to the individual survey targets in advance, and only those who gave their consent were surveyed. The purpose and content of the interviews were explained verbally to the subjects themselves, and their consent was confirmed by asking them to sign a consent form. The subjects were promised that the content of the survey form would not be used for any purpose other than this research, that they would remain strictly anonymous, and that their privacy would be respected in the Ph.D. dissertation and the journal article.

### Consent to publish

Not applicable. No images or videos of participants are used in the writing of this piece.

### Availability of data and materials

All data used in this study can be obtained from the first author upon a reasonable request.

### Authors contribution

TY and KM accounted for the idea. TY and SN collected the data. TY, AMM, and RE performed the data analysis, and TY, AMM, RE, EMM, NTH, and KM interpreted the data. All authors contributed to writing the manuscript and approved the final version before submission for publication.

## Data Availability

All relevant data are within the manuscript and its Supporting Information files.

## Acknowledgments

We would like to thank several people for their help and support in the preparation of this paper. Particularly, we would like to express our deepest gratitude to the elderly people of Kumejima-cho, Okinawa Prefecture, for their sincere cooperation in the survey. We would also like to express our gratitude to the Kumejima Town Council of Social Welfare for their kind assistance in the field. This study is based on part of the author’s doctoral thesis (2012) and has been reorganized with the addition of new findings. The author would like to express his sincere gratitude to the late Professor Shigeji Miyagi for his enthusiastic guidance during his doctoral thesis research. We sincerely pray for the repose of the soul of Professor Shigeji Miyagi.

## List of abbreviations

AGFI: adjusted goodness-of-fit index
AIC: Akaike information criterion
BMI: body mass index
CFI: comparative fit index
CROSS: Consensus-Based Checklist for Reporting of Survey Studies
GFI: goodness-of-fit index
QoL: quality of life
RMSEA: root mean square error of approximation
SP Health Scale: Spirituality Health Scale for the Elderly
WHO: World Health Organization

